# Environment-Wide Association Study of Cognitive Function in U.S. Older Adults using the NHANES Data

**DOI:** 10.1101/2024.07.22.24310659

**Authors:** HyunA Jang, Jiyun Lee, Vy Kim Nguyen, Hyeong-Moo Shin

## Abstract

Neurodegenerative diseases pose increasing challenges to global aging populations. Cognitive decline in older adults is an initial indicator of neurodegenerative diseases, yet comprehensive research on environmental chemical exposures related to cognitive decline is limited. This study uses Environment-Wide Association Study (EWAS) framework to investigate associations of environmental chemicals with cognitive function in individuals aged ≥60 years. We used the Digit Symbol Substitution Test (DSST) scores (lower scores indicate cognitive decline) and chemical biomarker data of the U.S. National Health and Nutrition Examination Survey (NHANES) spanning four cycles (1999-2000, 2001-2002, 2011-2012, 2013-2014). We conducted multiple survey-weighted regression to identify biomarkers associated with DSST scores, penalized logit regression to estimate odds ratio (OR) of cognitive decline with identified biomarkers, and correlation network analyses to examine relationships among identified biomarkers and cognitive decline. After correction for multiple comparisons, 27 out of 173 biomarkers having a ≥10% detection rate were associated with DSST scores (*q*-value <0.05). Among them, increased odds of cognitive decline were associated with elevated levels of blood lead (Pb) (OR = 1.12, 95% CI: 1.01,1.42), blood 1,4-dichlorobenzene (1,4-DCB) (OR = 1.34, 95% CI: 1.17, 1.54), and urinary 4-(methylnitrosamino)-1-(3-pyridyl)-1-butanol (NNAL) (OR = 1.34, 95% CI: 1.10, 1.62). Correlation network showed biomarkers that potentially impact cognitive decline upon related health conditions, such as stroke. In conclusion, leveraging the EWAS framework enables us to identify chemical biomarkers that were not previously discovered from traditional approaches of examining a small number of chemicals at a time. While our findings provide foundation for further research, longitudinal studies are warranted to elucidate causal relationships.

## Introduction

Increasing incidence of neurodegenerative diseases poses a prominent challenge in societies experiencing global population aging.^1^ In the United States (U.S.) alone, more than 6.2 million people were affected by neurodegenerative diseases in 2022, including Alzheimer’s disease.^2^ These diseases accounted for approximately 300,000 deaths and 3 million disability-adjusted life years (DALYs) during 1990-2016.^3–5^ The prevalence of clinical Alzheimer’s disease among adults aged 65 years and older is projected to increase from 7.2 million in 2025 to 13.9 million in 2060.^6^ In 2021, the U.S. incurred an annual cost of over $355 billion due to Alzheimer’s disease and related dementias, and this societal burden is expected to increase due to the increased health care cost.^7^ Consequently, there is growing attention towards the identification of risk factors and treatment development for neurodegenerative diseases.^8^

Neurodegenerative diseases are characterized by the progressive loss of neurons, leading to deterioration in both the structure and function of neural networks.^9^ This deterioration ultimately leads to impaired cognitive function.^1,10^ Gradual cognitive decline in old age is typically limited to subtle declines that evolve slowly over the years, attributed to normative developmental processes.^11,12^ Conversely, mild or precipitous declines in cognitive function can lead to pathological processes underlying Alzheimer’s disease and related dementias.^11,13^ Thus, early detection of changes in cognitive function, such as mild cognitive impairment, enables timely diagnosis and treatment of neurodegenerative diseases, potentially delaying disease progression.^13,14^

The risk of cognitive decline generally increases due to a combination of genetic and non-genetic factors. Age and genetic factors, including family history and susceptibility genes such as the apolipoprotein E ε4 allele, are the most significant contributors to cognitive decline.^15,16^ Other well-known non-genetic risk factors include comorbidities (hypertension, diabetes, stroke), unhealthy diet, physical inactivity, smoking, and alcohol consumption.^17,18^ These risk factors could influence the susceptibility of individual’s cognitive function (e.g., disease onset, timing, severity) by interacting with genetic factors.^16,19,20^ Therefore, identifying and understanding modifiable risk factors of cognitive decline could be a proactive approach to promoting cognitive health.^17,21^

Several studies have assessed associations between exposures to environmental chemicals and cognitive decline in specific populations. For example, exposures to metals, typically manganese (Mn),^22–24^ cadmium (Cd),^24–27^ lead (Pb),^25–27^ barium (Ba),^24^ cobalt (Co),^24^ cesium (Cs),^24^ and thallium (TI)^24^ were reported to increase the risk of low cognitive performance. In addition, higher urinary 3-phenoxybenzoic acid concentrations in adults were associated with cognitive dysfunction^28^ and it was observed that butyl benzyl phthalate is a potential cognitive-disrupting compound.^29^ However, previous studies examined exposure to a small number of chemicals or compounds individually in association with cognitive function. In addition, they are neither systematic nor comprehensive.

Environmental-Wide Association Studies (EWAS), derived from Genome-Wide Association Studies (GWAS), are a useful approach for systematically and comprehensively evaluating associations between hundreds of environmental factors and health outcomes.^30^ While GWAS aim to identify genetic factors associated with diseases of interest, EWAS focus on assessing environmental factors. Over the past decade, several EWAS studies have been conducted to elucidate the relationship between environmental chemical exposures and health outcomes.^31^ Frequently-studied health outcomes were type 2 diabetes,^30,32,33^ obesity,^34,35^ and blood pressure.^36,37^ Other outcomes included liver enzymes,^38^ mental and social well-being,^39^ testosterone deficiency,^40^ diabetic retinopathy,^41^ multiple sclerosis,^42^ peripheral arterial disease,^43^ and cardiovascular disease.^44^ However, no studies have simultaneously and comprehensively examined environmental chemical exposures associated with cognitive decline in older adults.

This study aims to identify environmental chemical biomarkers associated with cognitive decline in U.S. older adults by using the EWAS approach and the National Health and Nutrition Examination Survey (NHANES) data. First, to screen chemicals or their biomarkers, we examined a single association for each biomarker and identified those that were statistically significant after controlling for multiple comparisons. Second, we evaluated the effects of multiple exposures to those identified biomarkers on cognitive decline. Finally, we extracted a correlation network structure to explore potential pathways between chemical exposures and cognitive decline and further assessed the potential mediating effect of other modifiable factors.

## 2. Methods

### 2.1. Study population

NHANES is conducted every two years in the U.S. by the Centers for Disease Control and Prevention (CDC) and the National Centers for Health Statistics (NCHS). This is a cross-sectional survey designed to assess the health and nutritional status of both adults and children. To obtain a representative sample of the U.S. population, complex, multi-stage, and probability sampling techniques are used. NHANES data are publicly available (at https://www.cdc.gov/nchs/nhanes) and include individual questionnaires, physical examinations, and laboratory tests. More details on the design and methods of the NHANES data are available elsewhere.^45^

Our study combined data from four cross-sectional surveys conducted in 1999-2000, 2000-2001, 2011-2012, and 2013-2014, each of which included assessments of cognitive function. Targeting older adults (age ≥60 years), our analysis consisted of 4,970 individuals after excluding those with missing cognitive function data (n = 34,774) and those with abnormal urinary creatinine levels (n = 1,191) (Figure 1). The NCHS Ethics Review Board has approved this survey (Protocol #98-12 for NHANES 1999-2002 data, Protocol #2011-17 for NHANES 2011-2014 data), and all participants completed informed consent forms.

**Figure 1.**
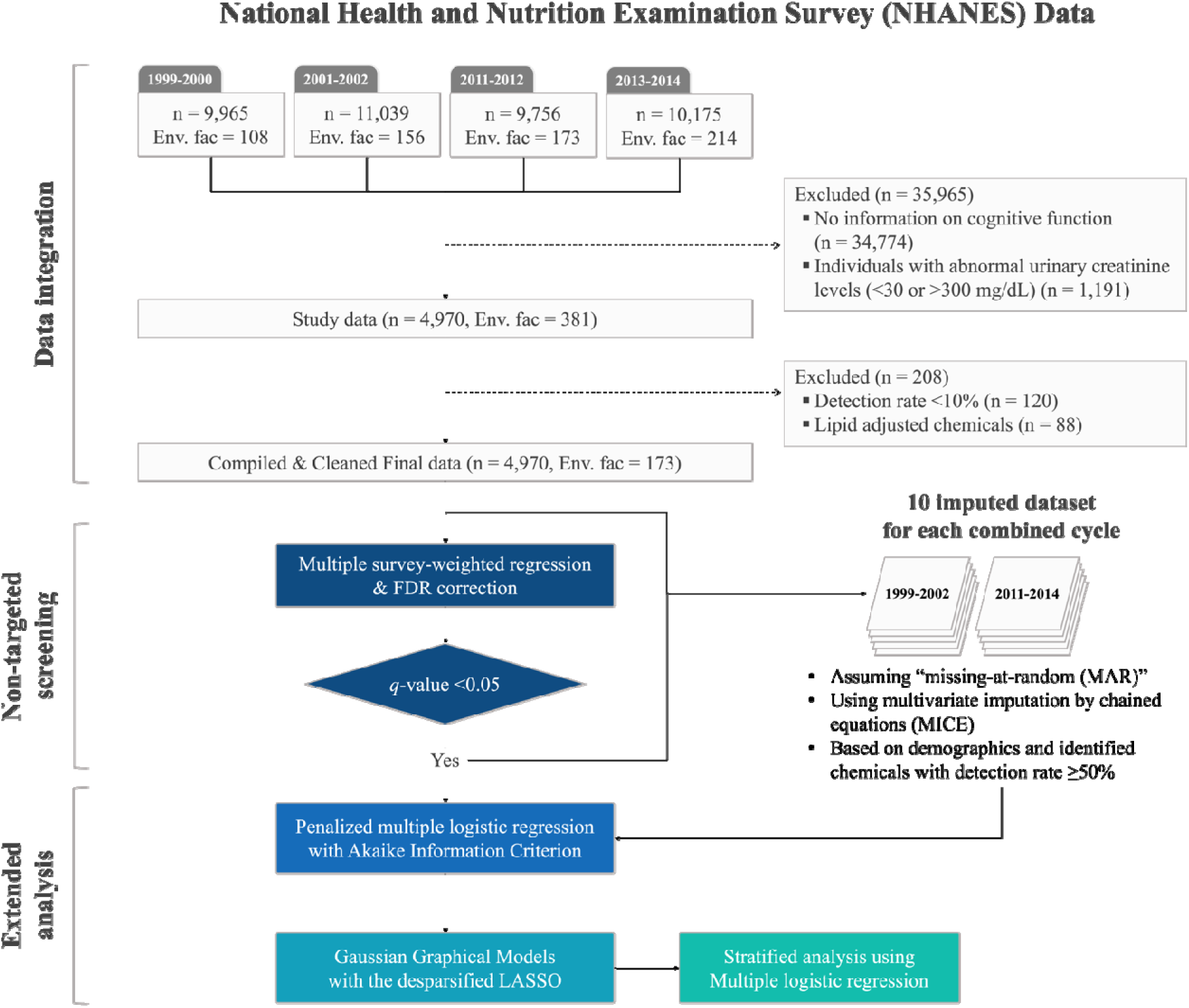
The process of data integration, non-targeted screening, and extended analysis. Data from four cycles of the National Health and Nutrition Examination Survey (NHANES) were used in this study. Final study subjects and environmental factors included in this study were selected based on specified inclusion and exclusion criteria.

### 2.2. Cognitive function assessment

In NHANES, the Digit Symbol Substitution Test (DSST), a subscale of the Wechsler Adult Intelligence Scale, was utilized to evaluate primary cognitive processing speed, attention, and working memory.^46^ Its application aids in determining the presence of cognitive dysfunction across various stages, including preclinical, prodromal, and dementia phases.^47,48^ Administered in a paper-and-pencil format, this cognitive test requires individuals to match symbols to corresponding numbers based on a provided key.^49^ The score is the total number of correct matches within 90 to 120 seconds, with lower scores indicating diminished cognitive function.^49^ Since 1999, NHANES has administered this test in participants aged 60 years and older. This test occurs during face-to-face interviews at the Mobile Examination Center by trained interviewers. Participants have the option to select from English, Spanish, Korean, Vietnamese, or Chinese. In this study, we used DSST z-scores when screening chemical biomarkers related to cognitive function and DSST raw scores below the 25^th^ percentile when defining as cognitive decline, as used by NCHS reports^50^ and other previous studies.^51,52^

### 2.3. Measurement of environmental chemical biomarkers

In NHANES, environmental chemical biomarker data were collected from the blood and urine samples of randomly selected participants within specific age groups in a one-third sample. Because detailed biological sample collection methods from NHANES participants vary by cycle,^53–56^ we used a harmonized and unified version of the NHANES chemical biomonitoring data.^57^ A total of 381 environmental chemical biomarkers were examined across the four cycles. These chemicals could be grouped as follows: acrylamide (n = 2), aldehydes (n = 13), aromatic amines (n = 16), dietary components (n = 6), dioxins (n = 14), furans (n = 20), metals (n = 29), per- and polyfluoroalkyl substances (PFAS) (n = 11), personal care & consumer product compounds (PCCPCs) (n = 12), pesticides (n = 55), phosphate flame retardants (PFR) (n = 9), phthalates & plasticizers (n = 14), polyaromatic hydrocarbons (PAHs) (n = 12), polychlorinated biphenyls (PCBs) (n = 70), smoking-related compounds (SRCs) (n = 15), and volatile organic compounds (VOCs) (n = 83). Detailed information, including the number of observations, detection rate, and lower limit of detection (LLOD) value, is given in Table S1 of Supplementary Information. In NHANES, all chemical biomarker concentrations below the LLOD were replaced with the LLOD/`√2`. The distribution of each biomarker concentration is provided in Table S2. The NHANES website offers detailed laboratory measurement methods and quality control procedures (https://wwwn.cdc.gov/nchs/nhanes/).

### 2.4. Covariates

We selected 11 covariates which are potential confounding or risk factors for cognitive decline identified in previous studies: sex (men and women), age group (60-64, 65-69, 70-74, 75-79, and ≥80 years), race/ethnicity (non-Hispanic white, non-Hispanic black, Mexican American/Hispanic, and others), marital status (married or living with partner, never married, divorced or separated, and widowed), education level (<high school, high school, and >high school), body mass index (<25, 25-<30, 30-<40, and ≥40 kg/m^2^),^58^ cotinine level as a proxy of smoking status (<3 and ≥3 ng/mL),^59^ alcohol consumption (non-drinker and drinker), health conditions (hypertension, diabetes, and stroke). Alcohol consumption and health conditions were ascertained by self-reported questionnaires (variable name in NHANES): “Had at least 12 alcohol drinks/lifetime? (ALQ110)”, “Ever told you had high blood pressure? (BPQ020)”, “Ever told you have diabetes? (DIQ010)” and “Ever told you had a stroke? (MCQ160f)”.

### 2.5. Statistical analysis

Descriptive statistics of participant characteristics were summarized using mean ± standard deviation (SD), number of cases (n), and percentage (%). The differences in DSST scores among each covariate were compared using t-tests and ANOVA. A cognitive decline group (i.e., participants with the lowest 25th percentile DSST scores) and a normal group (i.e., the rest as a reference) were compared using the chi-squared test. All analyses were performed using R version 4.3.1 (R Core Team, https://www.R-project.org/). The critical level of significance was set at α = 0.05.

#### 2.5.1. Screening environmental chemical biomarkers related to cognitive function scores

To screen hundreds of chemical biomarkers that are associated with lower DSST z-scores, we performed an EWAS non-targeted screening approach and identified biomarkers potentially affecting cognitive function. During this step, we used complete-cases data without missing values and considered chemicals with a detection rate of ≥10%. All concentrations were log10- transformed to control a right-skewed distribution and z-standardized. We explored the association between each biomarker and DSST z-scores using multiple survey-weighted regression adjusted for all covariates. To account for the complex survey design, we calculated sample weights following the NHANES guidelines.^60^ *P*-values were corrected using the false discovery rate (FDR, *q*-value) (Figure 1).

#### 2.5.2. Associations between exposure to identified biomarkers and cognitive decline

After identifying chemical biomarkers associated with DSST z-scores from the first step (*q*-value <0.05), we divided our participants into two groups (a normal group *vs*. a cognitive decline group) and examined associations of chemical exposures with cognitive decline by only including the identified chemical biomarkers with a detection rate of ≥50% and variance inflation factor (VIF) <10 in the model. We employed a multivariate imputation by chained equations (MICE) technique assuming ‘missing-at-random (MAR)’ to create 10 imputed datasets.^61^ To reduce imputation bias due to temporal trends in chemical biomarker, we generated imputed data separately for each of the 1999-2002 and 2011-2014 cycles and estimated the odds ratio (OR) and 95% confidence intervals (CI) of cognitive decline associated with biomarker levels using penalized logit regression with the Akaike Information Criterion (AIC) adjusted for all covariates.

#### 2.5.3. Interactive impact of chemical biomarkers and selected covariates on cognitive decline

To uncover the interactive impact of identified chemical biomarkers and the selected covariates on cognitive decline, we first combined imputed data from both the 1999-2002 and 2011-2014 cycles, focusing solely on chemical biomarkers consistently measured across all cycles. Next, we utilized Gaussian Graphical Models (GGMs) with the desparsified Lasso to derive a partial correlation network structure encompassing the identified chemical biomarkers, cognitive decline, and all covariates. Significant *p*-values were adjusted using the Holm– Bonferroni method. We selected non-chemical factors exhibiting statistically significant correlation coefficients with cognitive decline. Finally, we conducted stratified analyses on each category of the non-chemical factors to assess their potential as mediators using logistic regression adjusted for other covariates.

## 3. Results

### 3.1. Characteristics of study participants

Overall, 4,970 participants were included in this study, with 1,305 participants classified as the cognitive decline group. Significant differences in DSST z-scores were observed across all covariates (Table 1). Men had lower DSST z-scores than women, and Mexican Americans/Hispanics or widowed individuals had the lowest z-scores. DSST z-scores tended to decrease with increasing age or lower education levels. Smokers scored lower than non-smokers, whereas participants who consumed alcohol scored higher than those who never drank alcohol. No trend was observed with changes in BMI.

**Table 1.**
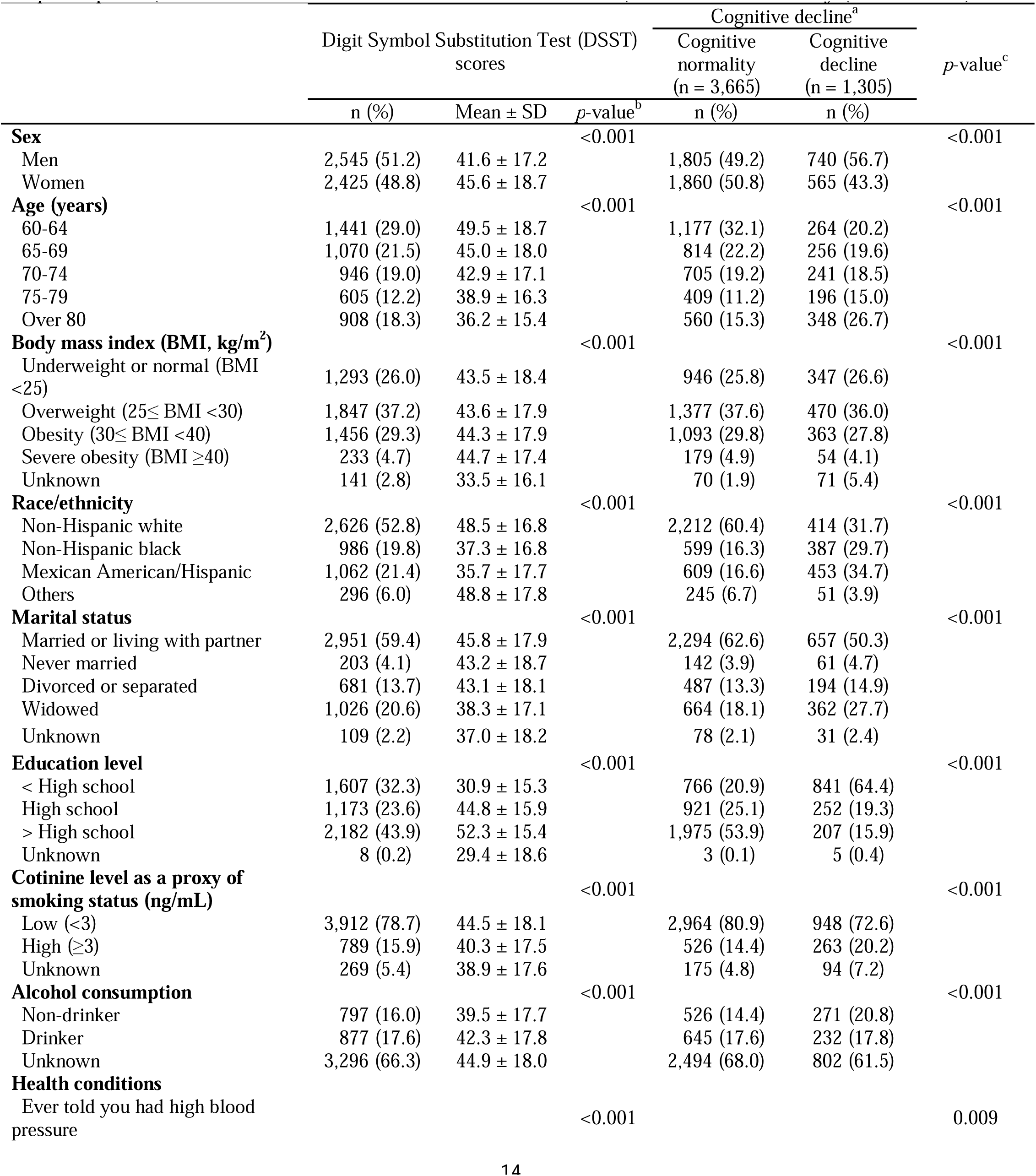
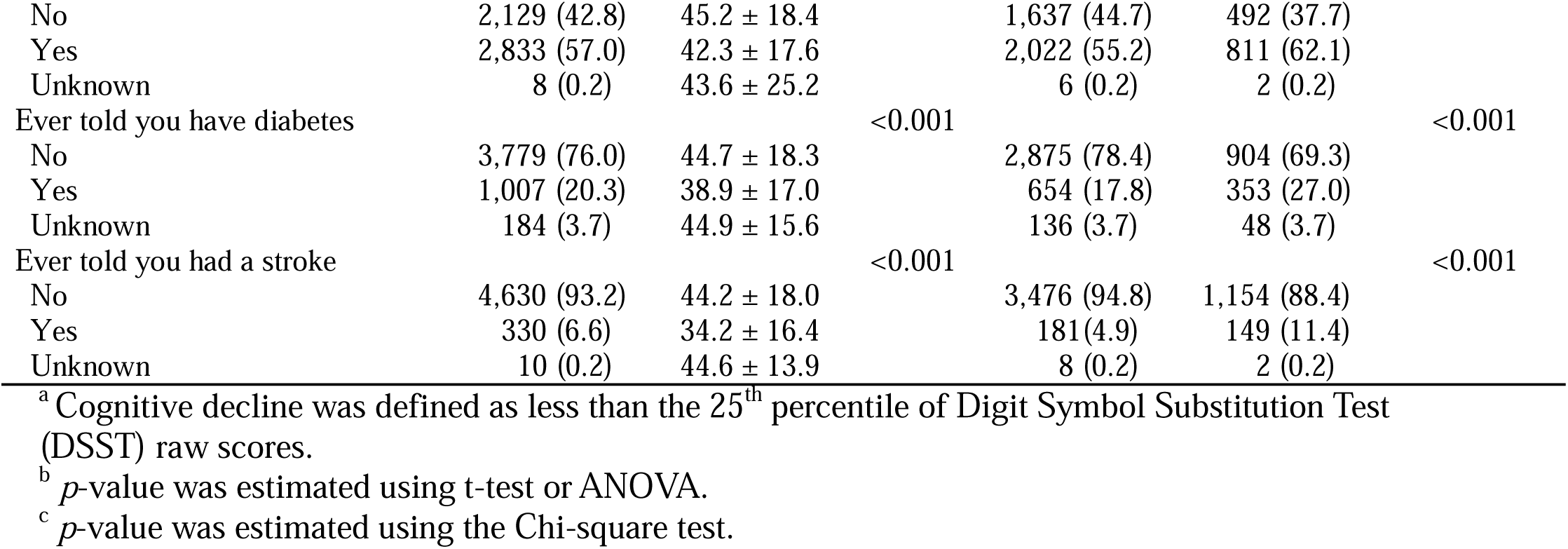
Characteristics of the National Health and Nutrition Examination Survey (NHANES) study articipants (1999-2000, 2001-2002, 2011-2012, and 2013-2014) included in this study (total = 4,970).

### 3.2. Identified chemical biomarkers related to cognitive function test scores

Out of 381 environmental chemical biomarkers examined across all four NHANES cycles, we only evaluated 173 biomarkers with a detection rate of ≥10% (Figure 1). Among the 173 chemicals, 27 were associated with DSST z-scores (*q*-value <0.05). These included metals (n = 11), PAHs (n = 3), PCBs (n = 3), pesticides (n = 3), phthalates (n = 2), and one each of PCCPCs, phytoestrogens, VOCs, flame retardants, and SRCs (Figure 2). PAHs, PCBs, phthalates, SRCs, and VOCs each decreased DSST z-scores, whereas flame retardants, PCCPCs, and phytoestrogens increased the scores. Metals and pesticides had varying effects depending on the biomarker (Table S3). For example, per 1 standard deviation increase in log10-transformed blood lead (Pb) level, DSST z-scores decreased by 0.09.

**Figure 2.**
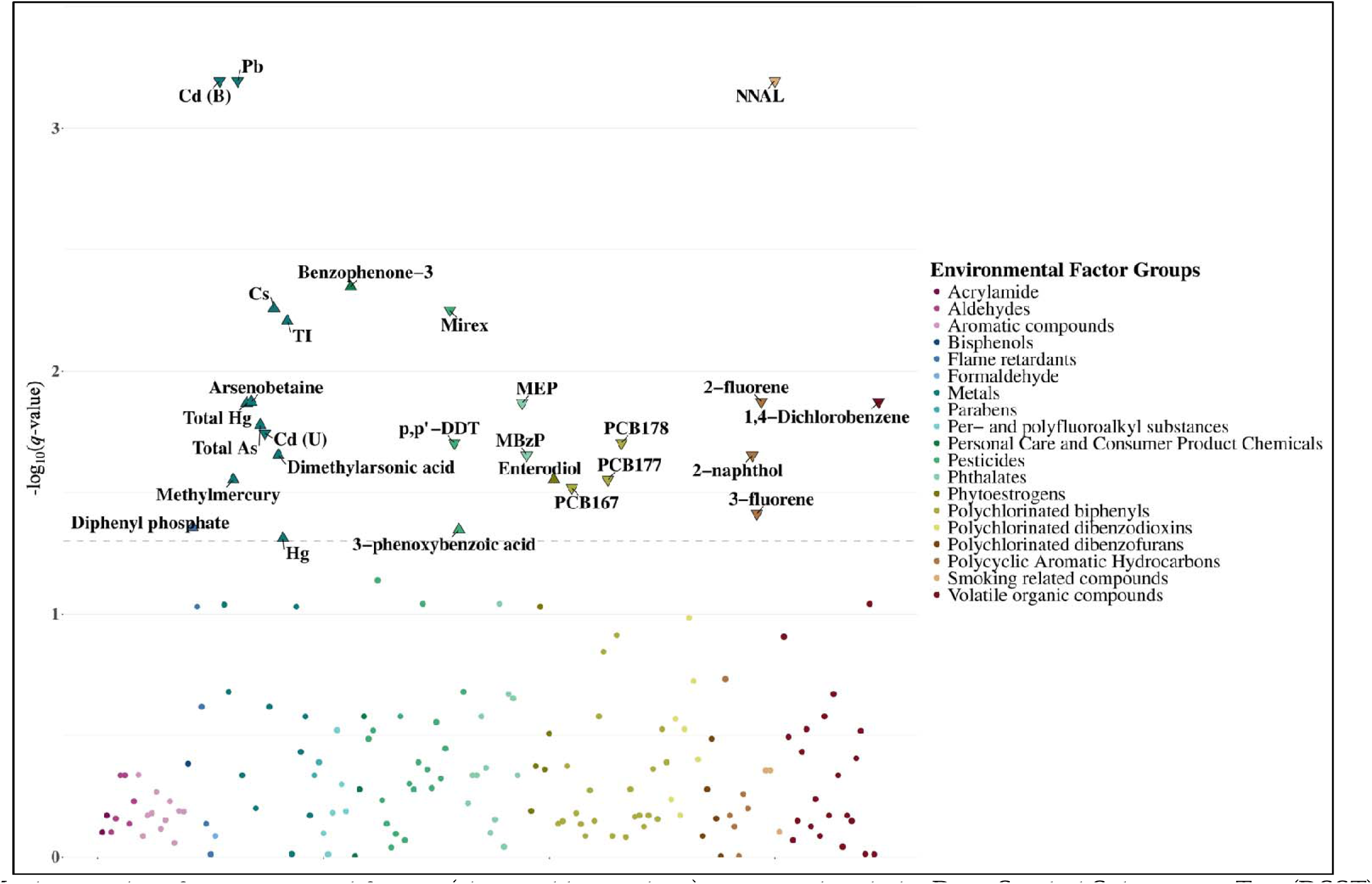
Manhattan plot of environmental factors (chemical biomarkers) associated with the Digit Symbol Substitution Test (DSST) z-scores. Significant chemical biomarkers were dotted above the grey dashed line (*q*-value <0.05), and the arrows indicated the direction of their effects on DSST z-scores. Abbreviation: B, blood; DDT, dichlorodiphenyltrichloroethane; NNAL, 4- (methylnitrosamino)-1-(3-pyridyl)-1-butanol; PCB, polychlorinated biphenyl; U, urine.

### 3.3. Associations between exposure to identified chemicals and cognitive decline

After screening chemical biomarkers that were associated with DSST z-scores, we examined associations between the identified biomarkers and cognitive decline by restricting analyses to those with a detection rate of ≥50% and separating the entire data to 1999-2002 and 2011-2014 cycles. When using the data collected during the 1999-2002 cycle, we could evaluate only 8 out of the 27 identified biomarkers, because the other 19 biomarkers were not measured during this cycle or had detection rates below 50%. From this cycle, blood Pb increased the odds of having cognitive decline (OR = 1.12, 95% CI: 1.01, 1.42), whereas blood cadmium (Cd) decreased the odds (OR = 0.78, 95% CI: 0.64, 0.96) (Table 2). From the 2011-2014 cycle data, we evaluated 18 biomarkers after excluding those that were not measured during this cycle or highly correlated chemicals with a VIF >10. From this cycle, blood 1,4-DCB and urinary 4- (methylnitrosamino)-1-(3-pyridyl)-1-butanol (NNAL) increased the odds of having cognitive decline (OR = 1.34, 95% CI: 1.17, 1.54 and OR = 1.34, 95% CI: 1.10, 1.62, respectively).

**Table 2.**
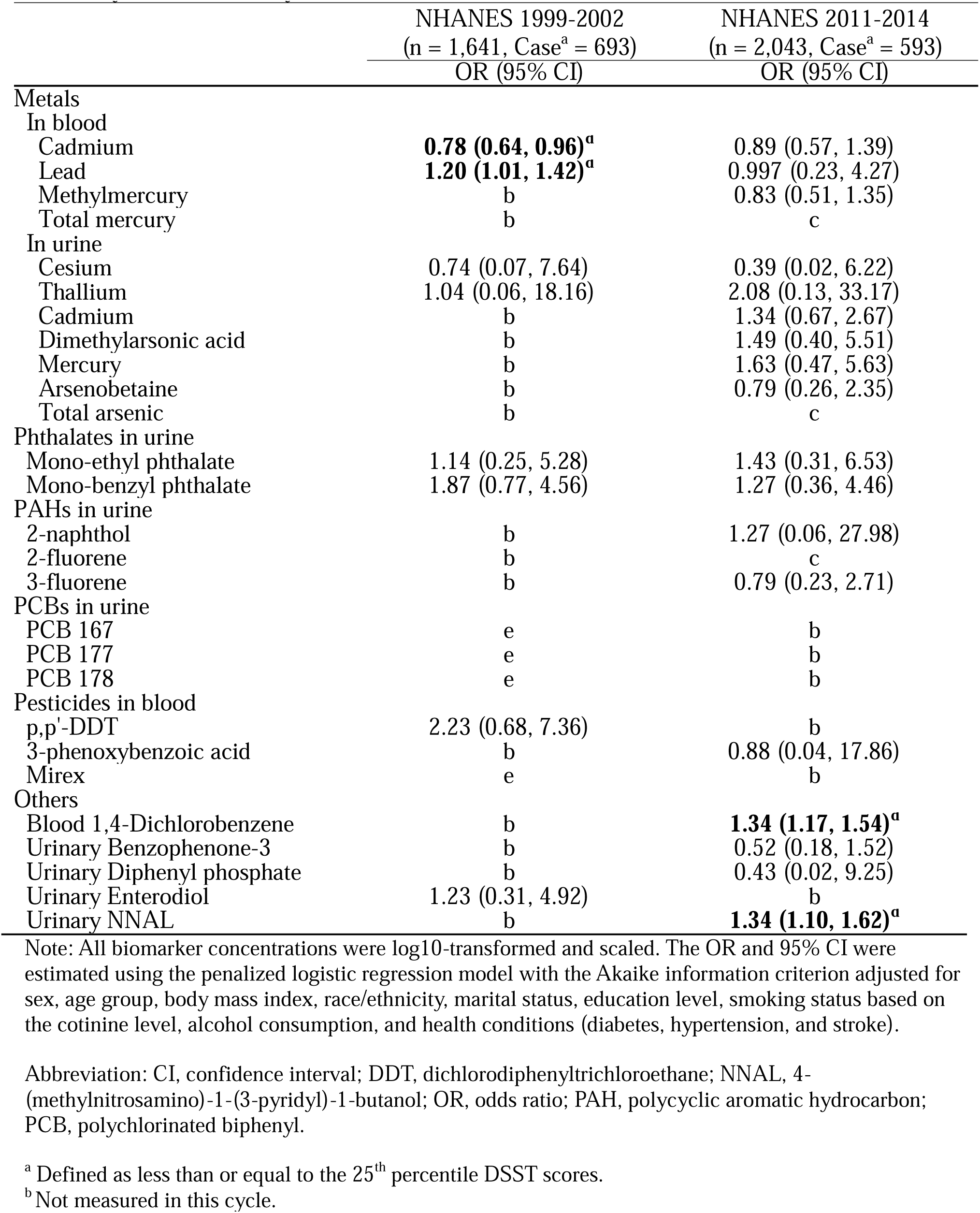
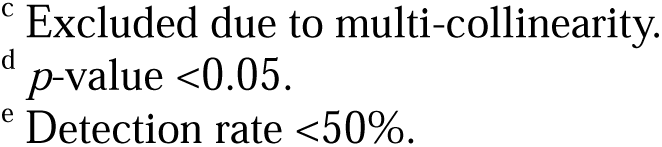
Associations between 27 identified environmental chemical biomarkers and cognitive decline by two combined cycles (NHANES 1999-2002 and 2011-2014) (total = 4,970).

|  | NHANES 1999-2002<br>(n = 1,641, Case<sup>a</sup> = 693) | NHANES 2011-2014<br>(n = 2,043, Case<sup>a</sup> = 593) |
| --- | --- | --- |
|  | OR (95% CI) | OR (95% CI) |
| Metals |  |  |
| In blood |  |  |
| Cadmium | **0.78 (0.64, 0.96)<sup>a</sup>** | 0.89 (0.57, 1.39) |
| Lead | **1.20 (1.01, 1.42)<sup>a</sup>** | 0.997 (0.23, 4.27) |
| Methylmercury | b | 0.83 (0.51, 1.35) |
| Total mercury | b | c |
| In urine |  |  |
| Cesium | 0.74 (0.07, 7.64) | 0.39 (0.02, 6.22) |
| Thallium | 1.04 (0.06, 18.16) | 2.08 (0.13, 33.17) |
| Cadmium | b | 1.34 (0.67, 2.67) |
| Dimethylarsonic acid | b | 1.49 (0.40, 5.51) |
| Mercury | b | 1.63 (0.47, 5.63) |
| Arsenobetaine | b | 0.79 (0.26, 2.35) |
| Total arsenic | b | c |
| Phthalates in urine |  |  |
| Mono-ethyl phthalate | 1.14 (0.25, 5.28) | 1.43 (0.31, 6.53) |
| Mono-benzyl phthalate | 1.87 (0.77, 4.56) | 1.27 (0.36, 4.46) |
| PAHs in urine |  |  |
| 2-naphthol | b | 1.27 (0.06, 27.98) |
| 2-fluorene | b | c |
| 3-fluorene | b | 0.79 (0.23, 2.71) |
| PCBs in urine |  |  |
| PCB 167 | e | b |
| PCB 177 | e | b |
| PCB 178 | e | b |
| Pesticides in blood |  |  |
| p,p'-DDT | 2.23 (0.68, 7.36) | b |
| 3-phenoxybenzoic acid | b | 0.88 (0.04, 17.86) |
| Mirex | e | b |
| Others |  |  |
| Blood 1,4-Dichlorobenzene | b | **1.34 (1.17, 1.54)<sup>a</sup>** |
| Urinary Benzophenone-3 | b | 0.52 (0.18, 1.52) |
| Urinary Diphenyl phosphate | b | 0.43 (0.02, 9.25) |
| Urinary Enterodiol | 1.23 (0.31, 4.92) | b |
| Urinary NNAL | b | **1.34 (1.10, 1.62)<sup>a</sup>** |

Note: All biomarker concentrations were log10-transformed and scaled. The OR and 95% CI were estimated using the penalized logistic regression model with the Akaike information criterion adjusted for sex, age group, body mass index, race/ethnicity, marital status, education level, smoking status based on the cotinine level, alcohol consumption, and health conditions (diabetes, hypertension, and stroke).
Abbreviation: CI, confidence interval; DDT, dichlorodiphenyltrichloroethane; NNAL, 4-(methylnitrosamino)-1-(3-pyridyl)-1-butanol; OR, odds ratio; PAH, polycyclic aromatic hydrocarbon; PCB, polychlorinated biphenyl.
<sup>a</sup> Defined as less than or equal to the 25<sup>th</sup> percentile DSST scores.
<sup>b</sup> Not measured in this cycle.
<sup>c</sup> Excluded due to multi-collinearity.
<sup>d</sup> $p$ -value <0.05.
<sup>e</sup> Detection rate <50%.

### 3.4. Interactive impact of chemical exposures and health conditions on cognitive decline

Based on the partial correlation network structure newly created in the current study, cognitive decline was directly associated with non-chemical factors, including smoking, obesity, alcohol consumption, and health conditions (stroke, diabetes, and hypertension) (Figure 3). Environmental chemical exposures seemed to have indirect relationships with cognitive decline, with health conditions serving as potential mediators. For instance, blood Cd was positively correlated with stroke which was also positively correlated with cognitive decline. Thus, we conducted stratified analyses by the status of stroke, diabetes, hypertension, and obesity to examine the potential interactive impact of health conditions. Overall, we observed no significant associations from these analyses (Figure 4). However, the odds of having cognitive decline for stroke cases tended to be higher than for non-cases with increased levels of blood Cd, blood Pb, urinary thallium (Tl), urinary mono-ethyl phthalate (MEP), and urinary mono-benzyl phthalate (MBzP). Similarly, the odds for diabetes cases tended to be higher than for non-cases with increased levels of blood Cd, urinary Tl, urinary cesium (Cs), urinary MEP, and urinary MBzP. The odds for hypertension cases tended to be higher than for non-cases with increased levels of urinary Tl. The odds for obesity cases tended to be higher than for non-cases with increased levels of urinary Tl, urinary MEP, and urinary MBzP.

**Figure 3.**
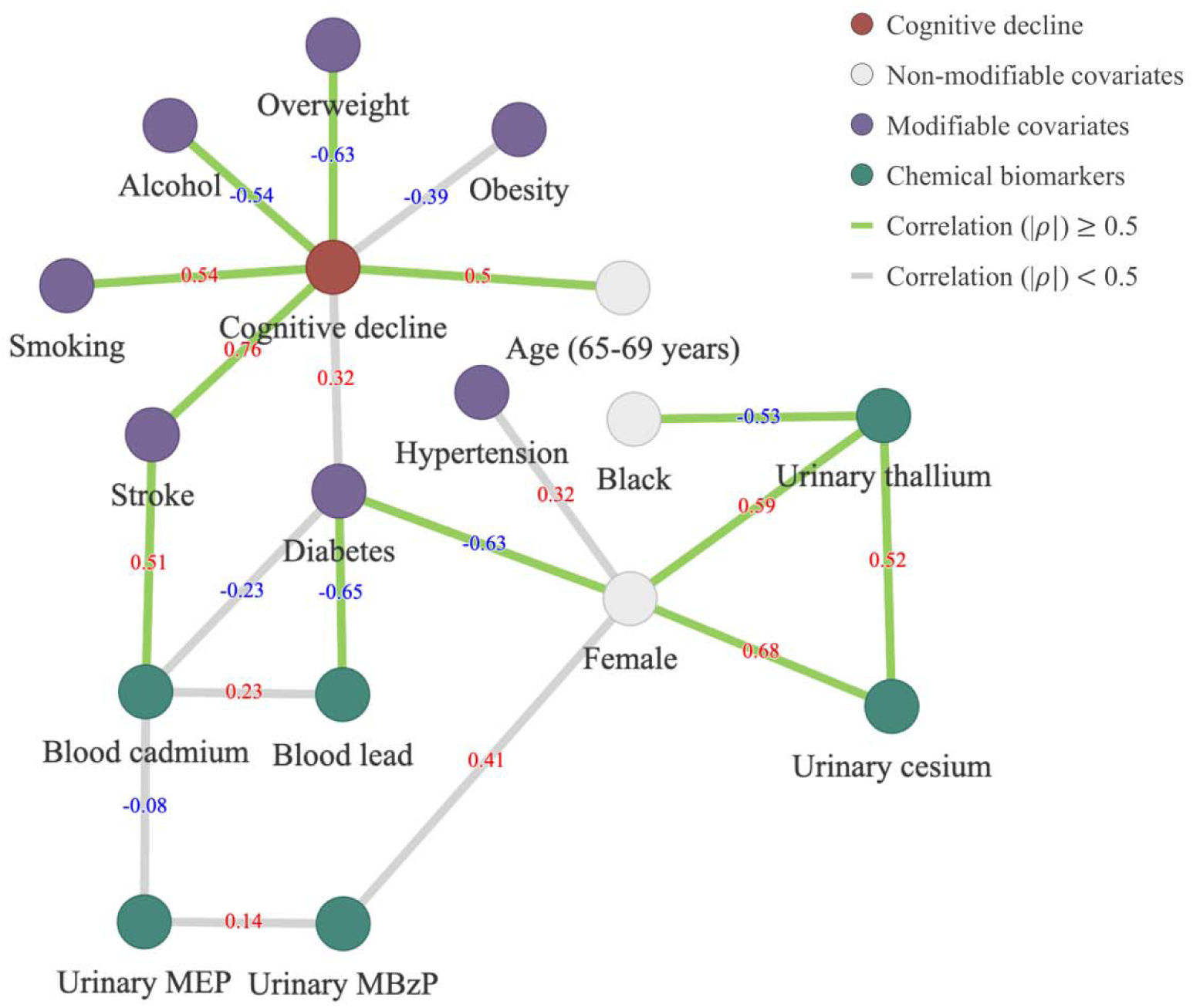
Partial correlation ( ) network plot depicting the magnitude and direction of correlation coefficients among identified chemical biomarkers, cognitive decline, and selected covariates. Only chemical biomarkers or covariates with significant correlation coefficients with cognitive decline (adjusted *p*-value <0.05) are shown. Green lines indicate relatively strong correlation coefficients (| 0.5). Red numbers indicate positive correlations and blue numbers indicate negative correlations.

**Figure 4.**
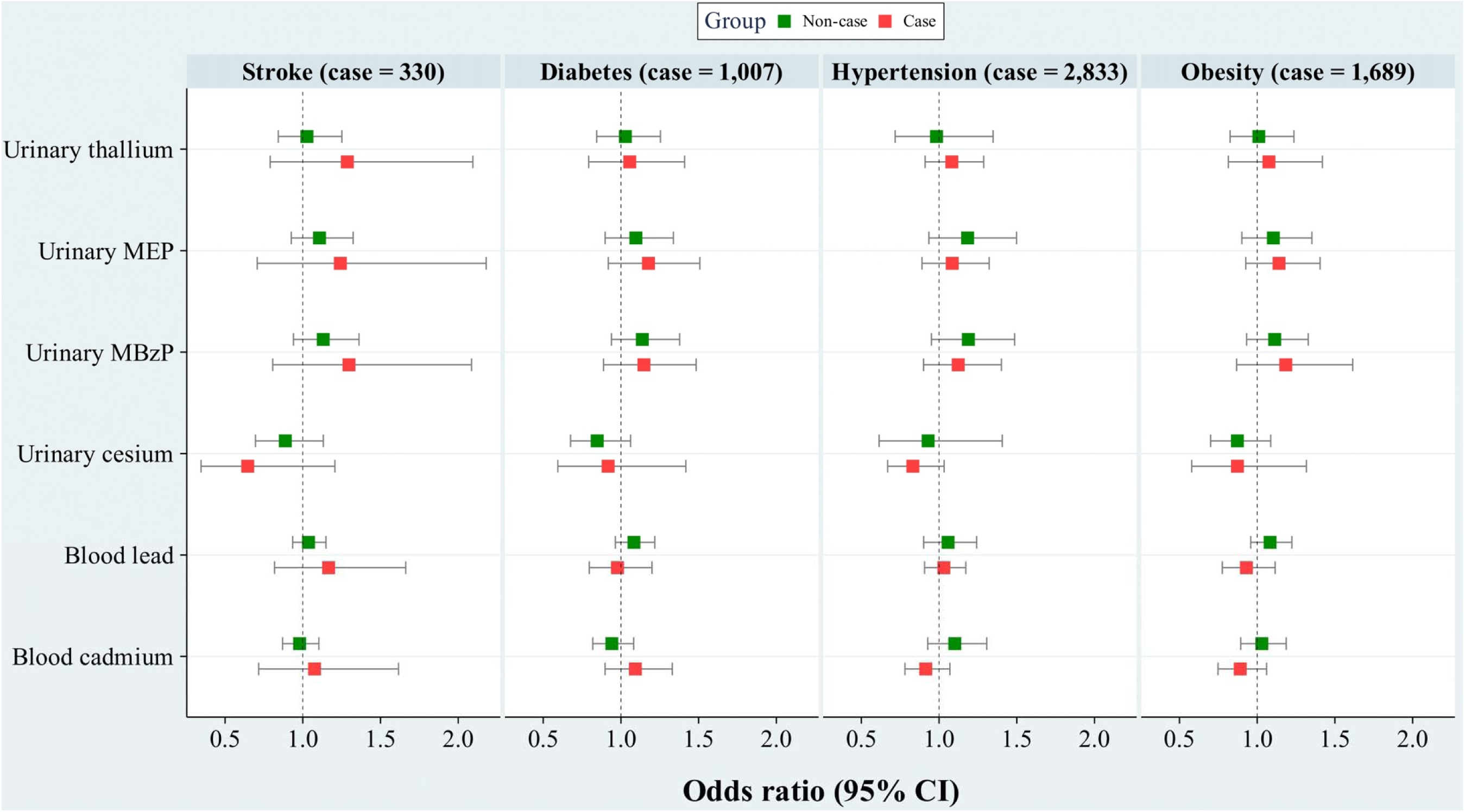
Odds ratio (OR) and 95% confidence interval (CI) of cognitive decline associated with six chemical biomarkers identified from correlation network structure stratified by four health conditions (total = 4,970). All chemical biomarker concentrations were log 10-transformed and scaled.

## 4. Discussion

To the best of our knowledge, this study represents comprehensive and systematic investigation into the association between environmental chemical exposures and cognitive decline. By using the EWAS approach and the NHANES data, we screened 173 environmental chemical biomarkers and identified those potentially related to cognitive decline. Additionally, we estimated the odds of having cognitive decline from the identified biomarkers and investigated previously undiscovered pathways linking chemical biomarkers to cognitive decline by extracting a correlation network structure. This allowed us to understand that cognitive decline can be worsened with increased chemical exposures among those with known risk factors of cognitive decline.

Among 173 chemical biomarkers investigated in this study, we observed that 27 biomarkers were associated with DSST z-scores. Most of them were metals, PAHs, PCBs, pesticides, and phthalates which are known to have neurotoxicity.^62–66^ Because these are characterized by persistence and bioaccumulation,^67–70^ older adults might have been chronically exposed throughout their lives.

When including all environmental chemical biomarkers identified from non-targeted screening, we found that blood Pb, urinary NNAL, and urinary 1,4-DCB increased the odds of cognitive decline. Previous studies have shown consistent results for the association between Pb exposure and cognitive function. For instance, each 1 µg/L increase in Pb concentration was associated with a 1.08-point decrease in DSST scores among U.S. older adults.^26^ For every 1 µg/dL increase in Pb levels, the odds of low cognitive performance in older adults in the U.S. by 10%.^25^ In addition, Pb, a ubiquitous environmental contaminant,^71^ is exposed primarily through food, water, tobacco smoke, air, dust, and soil.^72^ When Pb enters our human body, over 90% is accumulated in bones.^73^ Thus, older adults have high levels of Pb in their bodies due to their long lifespan.^73,74^ Pb can affect cognitive decline by disrupting neurotransmission^75^ and promoting oxidative stress^76^ through altering the fluidity or permeability of cell membranes.^77^ It also has indirect effects due to its associations with hypertension and other cardiovascular diseases,^78^ as well as its long-term storage in bones.^79^ Regarding NNAL, a previous study using NHANES data in older adults found that the group with the highest urinary NNAL concentration had a significantly lower DSST z-score by 0.19 compared to the lowest concentration group.^80^ NNAL is a reliable biomarker for nitrosamine ketone (NNK) exposure.^81^ NNK is present in the leaves of Nicotiana tabacum grown in the U.S.^82^ and can be exposed directly or indirectly through smoking,^83^ a well-known risk factor of cognitive decline. This compound is known to increase neurotoxicity by altering responses to centrally acting drugs, increasing susceptibility to neurotoxins, and enhancing metabolism.^81^ Lastly, 1,4-DCB (or paradichlorobenzene) can be easily encountered in our daily lives because it is an active ingredient in mothballs, deodorizers, and fumigants.^84,85^ However, there have been no epidemiological studies on the relationship between 1,4-DCB and cognitive function. Several *in vivo* studies have suggested its neurotoxic mechanism. This compound has central nervous system toxicity, causing oxidative damage and trace element alterations in various tissues, as well as oxidative DNA damage, leading to reported cognitive decline.^84,86,87^

From our correlation network analysis showing several pathways between chemical biomarkers and cognitive decline, we observed that stroke had relatively strong correlation coefficients with both blood Cd and cognitive decline. This finding highlights the need of this network analysis because blood Cd had protective effects against cognitive decline when considering other identified environmental chemicals in the regression. Previous studies have shown that exposure to Cd can impair cognitive performance both in isolated exposure scenarios and in combination with other environmental chemicals identified through EWAS screening.^24–27^ It is also well known that Cd causes vascular damage and promotes atherosclerosis^88^ by generating reactive oxygen species, disrupting sulfhydryl homeostasis, and downregulating nitric oxide.^89^ Stroke affects cognitive domains such as attention, memory, and language. People who had stroke commonly experience cognitive impairment and memory loss, making them highly likely to develop dementia within one year of stroke onset.^90,91^ While our findings suggest that the effects of Cd exposure on cognitive decline may vary by individual’s health conditions, the specific mechanisms underlying these pathways remain unclear. Further experimental investigation is warranted to elucidate the mediating effects of these health conditions on the association between Cd exposure and cognitive decline.

Our study has several limitations. Firstly, the cross-sectional nature of the NHANES data precludes establishing causation from our findings. Secondly, chemical biomarker concentration data were from a spot sample of each participant, so they may not represent individual’s average exposure over the lifetime of our participants. Thirdly, despite adjusting for confounders associated with cognitive decline based on previous studies, there may still be unmeasured confounding variables influencing our effect estimates. Fourthly, there could be imputation bias on the biomarker data because each NHANES cycle had a different list of chemical biomarkers and only one third of samples from each cycle were used to assess chemical biomarker concentrations. In addition, if the missing data mechanism of NHANES is non-random (i.e., missing not at random, MNAR), performing multiple imputation under the assumption of missing at random (MAR) may produce biased estimates.^92^ Finally, the EWAS approach used in this study cannot assess the effects of chemical mixtures, such as additive effects, synergism, potentiation, and antagonism.

## 5. Conclusions

In this study, we employed an EWAS approach to explore 173 environmental chemical biomarkers in association with cognitive decline among U.S. older adults. From the regression including all identified biomarkers significantly associated with cognitive decline, we observed positive associations between cognitive decline and Pb, 1,4-DCB, and NNAL. From the correlation network structure, Cd may increase the risk of stroke which is a risk factor of cognitive decline. These findings suggest that combined and prolonged exposure to these chemicals may worsen cognitive impairment in aging populations. Our study underscores the complex interplay between exposure to environmental chemicals and cognitive function in older adults, emphasizing the need for further research to elucidate causal mechanisms and effectively guide public health interventions.

## Author contributions

**HyunA Jang:** conceptualization, data curation, formal analysis, methodology, software, visualization, and writing – original draft. **Jiyun Lee:** conceptualization, data curation, methodology, visualization, validation, and writing – review & editing. **Vy Kim Nguyen:** resources, data curation, and writing – review & editing. **Hyeong-Moo Shin:** conceptualization, supervision, validation, and writing – review & editing.

## Data availability

The data and R code can be shared upon request to the corresponding author.

